# The Status of Indian Cancer Record Keeping and Study of Mesothelioma Cases to Ascertain Asbestos Exposure in India

**DOI:** 10.1101/2023.12.19.23300114

**Authors:** Raja Singh, Arthur L. Frank

## Abstract

**Setting:** Asbestos exposure causes mesothelioma which is classified as a malignancy and recorded by cancer registries which currently covers only16% of the population in India. The accurate number of mesothelioma cases may not be available for decision making as India still uses asbestos and it is important to ascertain its health impact, especially mesothelioma.

**Objective:** This study aims to find the cases of mesothelioma from 83 hospitals across India from 2012 onwards till 2022-2023. The study also compares the national registry reported cases to the ones found in this study.

**Design:** The study uses the Right to Information Act 2005 to find data from various hospitals and reporting the same. The data from the voluntary national registry was also collected. This was compiled and compared.

**Results:** Overall, the study shows 2213 cases of mesothelioma 2012 onwards in India from 83 hospitals. In the comparison period of 2012-2016, the registry reported cases were 54, while the study shows 1126 cases. Only 21% of hospitals in this study were part of the national registry programme.

**Conclusion:** Mesothelioma cases in India are more frequent than reported and the current recordkeeping for all cancers does not fully cover the expanse of India and needs to be revamped.

## Introduction

Mesothelioma is a malignancy which is intimately associated with exposure to asbestos^1–4^. It may occur in the pleura or peritoneum, with the latter being less frequent^5^. A study of the estimation of the global burden of mesothelioma deaths from incomplete national mortality data, came out with the figure of 38400 global mesothelioma related deaths yearly^6^. Many academic literature sources, including a case report from India published in 2009, stated that malignant pleural mesothelioma ‘is a rare tumor in the Western World and still rarer in India.^7^’This opinion, and the general small number of cases in percentage as compared to other cancers, also classified malignant mesothelioma into the rare category for India. ^8^ In a case report from an Indian physician, in 2019 it is stated that ‘there is no report from India of mesothelioma related to asbestos’ and the same paper discusses a 42 year old man who died due to pleural mesothelioma with enough evidence of prior asbestos exposure ^9^. This case report quoted the health statistics and health information systems of the World Health Organisation Mortality database of 2018. This reporting may be due to the smaller numbers, as compared to other cancers, but this rarity cannot be considered as absence of mesothelioma in India. On the other hand, in another study, which is stated by its authors to be the ‘largest study from India’ on malignant mesothelioma, the proven cases from 2015 to 2019 in the archives of the Department of Onco-pathology at Gujarat Cancer Research Institute were 128 ^10^.

This lack of consistency can be attributed to the unresolved issues in Indian cancer recordkeeping. Some sources have stated that there is a ‘very poor, almost non-existent, system to record death and disease’ in India, in addition to cancer being a non-notifiable disease in India^11^. As far as recordkeeping of cancer is concerned in India, there is a national cancer registry programme which started in December 1981 under the leadership of the Indian Council of Medical Research. But the total coverage of the population that is under the national cancer registry, as per the most recent on record reply by the department of health research in India, is 16%, though the same number is reported as 10% by the 139^th^ Report on Department Related Parliamentary Standing Committee on Health and Family Welfare on Cancer Care Plan & Management, Prevention, Diagnosis, Research & Affordability of Cancer treatment^12,13^. This low coverage of the population is something the committee expressed ‘its deep displeasure about’ and the committee ‘strongly’ believed that ‘there is an urgent need to have more rural PBCRs to get realistic information about the incidence and type of cancers across the country.’ The cancer registry programme in India is run by the National Centre for Disease Informatics Research (NCDIR), Bengaluru and has two types of registry systems ^14^. First is the Hospital Based Cancer Registries or HBCRs and the second is the Population Based Cancer Registries or PBCRs. The hospital-based registries have been set up in tertiary care centres with large capacity and are usually dedicated centres for cancer. Some of these centres also take up the record keeping work for hospitals in the same immediate geography, and these also operate Population Based Cancer Registries. As per the communication sent by NCDIR to the Ministry of Health and Family Welfare in July 2022, there are currently 38 PBCRs in the country covering both urban and rural areas, but the last published report with data is from data till 2016 and while the number of PBCRs at that time was 36, the report included data from only 28 as these PBCRs provided data which was ‘complete and met the desired quality ^15,16^.’ It is important to note that PBCRs collect data from multiple sources such as various clinics in multiple hospitals of various specialties, palliative care units, diagnostic centres as well as mortality data from death record centres ^16^. The data from 2016 onwards is still under process and is not available from NCDIR and due to the nature of agreement, the data is not available for policy, research, or epidemiological purposes unless published as a report in the public domain ^17^.

In order to study the actual trend of mesothelioma, especially since data post 2016 was not available, and because multiple hospitals were not under the coverage of NCDIR, this study was undertaken to ascertain a more accurate number of mesothelioma cases in India.

## Material and Methods

In the current study, the data from 83 hospitals has been collected from across the country using the Right to Information Act 2005^18,19^ or RTI Act, hereinafter. Hospitals were requested for the count of the diagnosed cases year wise from 2012 till 2022 and some extended till 2023. The applications were specifically made for data on mesothelioma which is classified under ICD-10 (of the International Classification of Diseases) as C45 ^20^. The period of collection of information was mid 2023.

The applications sent were provided with replies by the hospitals which were essentially public in nature, either being under the state government or the Central government or were substantially funded by these. This is because the method of collecting information, under the RTI Act is valid only for ‘public authorities’ under the RTI Act ^18^. Some replies needed further information and were appealed under the act and the final data received from all the hospitals was collated and collected.

Not all the 83 hospitals were under the PBCR, and this means that the study includes those hospitals which are otherwise not part of an organized cancer registry, which brings in data which has not been previously reported. The collected data was arranged and presented in this study to provide insights into the actual measurement of mesothelioma in India and brings fresh light to this disease which is almost always caused exclusively in people having previous exposure to asbestos, either occupational or in non-occupational settings.

The comparison was also made between the data collected in this study and the data available in the NCDIR report for a comparable period of 2012 to 2016, as this is the time period for which the NCDIR report was available ^16^. The comparison was with respect to the number of cases as well as the hospitals that were covered by the NCDIR database.

The special advantage of the Right to Information Act, 2005 is that data provided by the hospital under this statute of the Indian parliament is by default non personal, and its release makes it in the public domain. Therefore, this study is simply a collection of disease counts available in the public domain, with no personal detail of any patient, and no involvement of any human subject, which makes the study not under the purview of ethics approval.

There are 525 centres listed for treatments related to cancer under the radiation license by the atomic regulatory board in India ^21^. For the broad calculation of the number of hospitals, government or private providing cancer treatment, this may be considered as the universe size. According to this, 83 hospitals represent a sample size with a confidence level of 95% ±10% margin of error.

## Results

This study set out to find a more accurate number of mesothelioma cases in India. From the 83 hospitals under study, the total number of mesotheliomas from 2012 onwards till 2022, and in some cases till 2023, was 2213 cases. The list of hospitals that have been covered in the study are listed as Table 1.

**Table 1:** The list of hospitals state-wise of which data has been included in the study.

| S. No | Name of Hospital Included in this study | State |
| --- | --- | --- |
| 1 | Homi Bhabha Cancer Hospital & Research Centre, Visakhapatnam | Andhra Pradesh |
| 2 | Sri Venkateswara Institute of Medical Sciences, Tirupati | Andhra Pradesh |
| 3 | Govt. Hospital for Chest and Communicable Diseases, Guntur | Andhra Pradesh |
| 4 | Dr Bhubaneshwar Barooah Cancer Institute Guwahati | Assam |
| 5 | P.M.C.H. Patna | Bihar |
| 6 | Homi Bhabha Cancer Hospital and Research Centre, Muzaffarpur | Bihar |
| 7 | All India Institute of Medical Sciences Raipur | Chhattisgarh |
| 8 | Dr. Bhimrao Ambedkar Memorial Hospital, Raipur | Chhattisgarh |
| 9 | Goa Medical College | Goa |
| 10 | Guru Gobind Singh Hospital, Jamnagar | Gujarat |
| 11 | Department of Radiation Oncology Medical College & SSGH Vadodra | Gujarat |
| 12 | Gujarat Cancer and Research Centre GCRI Ahmedabad | Gujarat |
| 13 | GMERS Medical College and Hospital Navsari | Gujarat |
| 14 | Sardar Vallabhai Patel Institute of Medical and Research, Ahmedabad | Gujarat |
| 15 | Sir Takht Singhji Hospital, Bhavnagar | Gujarat |
| 16 | National Cancer Institute AIIMS Jhajjar | Haryana |
| 17 | Pt. BD Sharma PGIMS Rohtak | Haryana |
| 18 | Dr. Rajendra Prasad Government Medical College, Tanda | Himachal Pradesh |
| 19 | Indira Gandhi Medical College, Shimla | Himachal Pradesh |
| 20 | Bokaro Steel Plant Hospital | Jharkhand |
| 21 | Karnataka Institute of medical Sciences Hubli | Karnataka |
| 22 | KR Medical Mysore | Karnataka |
| 23 | Kidwai Memorial Institute of Oncology, Bengaluru | Karnataka |
| 24 | Malabar Cancer Centre | Kerala |
| 25 | T.D. Medical College Hospital Alappuzha | Kerala |
| 26 | General Hospital Ernakulam | Kerala |
| 27 | Govt medical college hospital, Kannur | Kerala |
| 28 | Govt medical college hospital, Kollam | Kerala |
| 29 | Govt Medical College Hospital, Kottayam | Kerala |
| 30 | Bhopal Memorial Hospital and Research Centre | Madhya Pradesh |
| 31 | AIIMS Bhopal | Madhya Pradesh |
| 32 | State Cancer Institute NSCB Medical College Jabalpur | Madhya Pradesh |
| 33 | Shyam Shah Medical College, Rewa | Madhya Pradesh |
| 34 | Tata Memorial Hospital | Maharashtra |
| 35 | ESIC ODC Mumbai | Maharashtra |
| 36 | Department of Radiation Oncology Cama & Albless Hospital Mumbai | Maharashtra |
| 37 | Bhabha Atomic Research Centre (Hospital) | Maharashtra |
| 38 | North Eastern Indira Gandhi Regional Institute of Health and Medical Sciences Shilong | Meghalaya |
| 39 | Rajinder Hospital, Patiala | Punjab |
| 40 | Homi Bhabha Cancer Hospital, Sangrur | Punjab |
| 41 | Homi Bhabha Cancer Hospital and Research Centre, Mullanpur | Punjab |
| 42 | Sawai Man Singh Hospital , Jaipur | Rajasthan |
| 43 | DR. S. N. Medical College & A. G. H. Mathuradas Mathur Jodhpur | Rajasthan |
| 44 | Govt Medical Hospital, Kota | Rajasthan |
| 45 | Acharya Tulsi Cancer Hospital and Research Institute | Rajasthan |
| 46 | TNCRP- Govt. Rajiv Gandhi General Hospital, Chennai | Tamil Nadu |
| 47 | TNCRP- Government Royapettah Hospital, Chennai | Tamil Nadu |
| 48 | TNCRP- Government Stanley Medical College, Chennai | Tamil Nadu |
| 49 | TNCRP- Institute of Obstetrics and Gynaecology, Chennai | Tamil Nadu |
| 50 | TNCRP- Madras Cancer Care Foundation, Chennai (Kumaran Hospital) | Tamil Nadu |
| 51 | TNCRP- The Tamil Nadu Government Multi Super Speciality Hospital, Chennai | Tamil Nadu |
| 52 | TNCRP- Apollo Cancer Centre | Tamil Nadu |
| 53 | TNCRP- Sri Ramachandra Medical College and Hospital | Tamil Nadu |
| 54 | TNCRP- Govt. Arignar Anna Memorial Cancer Hospital & Research Institute, Kanchipuram | Tamil Nadu |
| 55 | TNCRP- Govt. Viluppuram Medical College and Hospital | Tamil Nadu |
| 56 | TNCRP- Kovai Medical Centre and Hospital, Coimbatore | Tamil Nadu |
| 57 | TNCRP- G Kuppuswamy Naidu Memorial Hospital, Coimbatore | Tamil Nadu |
| 58 | TNCRP- Erode Cancer Centre, Thindal, Erode | Tamil Nadu |
| 59 | TNCRP- Thanjavur Cancer Centre, Thanjavur | Tamil Nadu |
| 60 | TNCRP- Govt Tirunelveli Medical College and Hospital, Tirunelveli | Tamil Nadu |
| 61 | TNCRP- International Cancer Centre, Neyyoor | Tamil Nadu |
| 62 | TNCRP- JIPMER Regional Cancer Centre, Puducherry | Tamil Nadu |
| 63 | Command Hospital Chennai | Tamil Nadu |
| 64 | Coimbatore Medical College Hospital | Tamil Nadu |
| 65 | Kalyan Singh Super Speciality Cancer Institute Lucknow | Uttar Pradesh |
| 66 | Institute of Medical Sciences BHU | Uttar Pradesh |
| 67 | Mahamana Pandit Madan Mohan Malviya Cancer Centre, Varanasi | Uttar Pradesh |
| 68 | Jawaharlal Medical College, AMU, Aligarh | Uttar Pradesh |
| 69 | LLRM Medical College, Meerut | Uttar Pradesh |
| 70 | Chittaranjan National cancer institute | West Bengal |
| **71** | Command Hospital | West Bengal |
| **72** | ESIC Joka | West Bengal |
| **73** | Government Medical College and Hospital | UT-Chandigarh |
| **74** | Nehru Hospital Postgraduate Institute of Medical education and Research | UT-Chandigarh |
| **75** | Lady Hardinge Medical College & Smt. S.K. Hospittal | UT-Delhi |
| **76** | Vardhaman Mahaveer Medical College and Safdarjung Hospital | UT-Delhi |
| **77** | Delhi State Cancer Institute | UT-Delhi |
| **78** | Kalavati Saran Childrens hospital KSC | UT-Delhi |
| **79** | Br Ambedkar Rotary Cancer centre, AIIMS Delhi | UT-Delhi |
| **80** | National Institute of Tuberculosis and Lung Diseases | UT-Delhi |
| **81** | Lok Nayak Hospital | UT-Delhi |
| **82** | Government Medical College Department of Radiation Oncology, Jammu | UT-J&K |
| **83** | Govt Medical College, Karanagar, Srinagar | UT-J&K |

The comparison was also made between the cases registered by the Indian National Cancer Registry Programme (by NCDIR) between 2012 to 2016 and the cases recorded in this study ^16^. While the NCDIR recorded 54 cases, this study recorded 1126 cases. The reasons for the difference in cases along with the limitations in the NCDIR report as well as this study have been discussed below. But it is important to note that out of the 83 hospitals that have been covered in the study, only 10 have a Hospital Based Cancer Registry under the NCDIR programme. Apart from this, only 21 (25%) out of the 83 are part of a Population Based Cancer Registry Programme of the NCDIR, This means that 62 (∼75%) of the hospitals are not part of the formal Indian record keeping mechanism under the Population National Cancer Registry Programme and 73 (∼88%) of the hospitals don’t have a hospital based registry under the programme,

In the data collection, one state in India, Tamil Nadu, had a state-based cancer registry which had 17 hospitals under its fold ^22^. For the state of Tamil Nadu, the hospitals under the registry have not been separately added, but the total registry numbers have been obtained. The other hospitals from Tamil Nadu, not part of the state registry have been added directly in this study.

## Discussion

It must be noted that the state of the Cancer Registry programme in India is dismal ^23^. Its coverage, with respect to the number of hospitals that remain uncovered, is not appropriate for a programme which started in 1981-82. Part of the reasons speculated for this may be the voluntary nature of inclusion in the database, as health is a state subject in India and it is the decision of individual states to include themselves into a national programme, or not. This could have been made mandatory, rather than the current voluntary inclusion, by making cancer a notifiable disease, as was recommended by the parliamentary standing committee ^13^. It is pertinent to note that many states have at the state level made cancer notifiable, and one state, Tamil Nadu, has a very robust cancer registry system as well. But at the national level, the Department of Health Research has turned down the suggestion of the parliamentary committee to make cancer notifiable by stating that cancer is a non-communicable disease and not an infectious disease which does not have community spread and ‘in the present circumstances, it may not be declared as notifiable disease ^12^.’ There are certain states in India such Andhra Pradesh, Chhattisgarh, Goa, Haryana, Himachal Pradesh, Jharkhand, Odisha and Rajasthan, which as per the 2022 communication of NCDIR do not have a Population Based Cancer Registry in India ^15^. This is along with Union Territories like Andaman Nicobar Islands, Chandigarh. Dadra and Nagar Haveli, Daman and Diu, Ladakh, Puducherry, and Lakshadweep. However, it is noted that major hospitals in some states without a PBCR are listed in the PBCR of an adjoining state. It must also be noted that the state level cancer data, even if fully calculated, would not provide the complete picture as people move from one state to another more developed state for treatment. An example of this is Gujarat, which has the Gujarat Cancer and Research Institute (GCRI). The total mesothelioma cases recorded at GCRI from 2012 to 2023 (July) have been 303, out of which 99 (∼32.67%) cases are from the adjoining Rajasthan and 23 (7.59%) from adjoining Madhya Pradesh. There are an additional 6 (∼1.99%) which are from other states and the total case load from Gujarat is 175 (∼57.75%). This shows the catchment area of certain established hospitals transcend beyond state boundaries, and the records from these states in that case may not be linked to the causative factors in those states. Additionally, the data from hospitals in this study are government or public hospitals, as the RTI Act only enables information from ‘public authorities’ and this excluded multiple private hospitals. This documents that more cases that would have been reported if private hospitals, not included in existing PBCRs, but with cancer facilities, would have been included. The exception to this is the Tamil Nadu Cancer Registry Project which has private hospitals data included and this study has taken the Tamil Nadu combined registry data instead of the breakdown from individual hospitals. The issue stated before of the national registry having limited coverage is also serious as this reporting is the one that is submitted to the International Cancer Registry and this becomes the basis of global policy decisions ^24^. Due to its limited coverage, the Indian database does not provide a complete picture of the scenario in India. Apart from that, in the experience of the authors, the NCDIR had missing data that was not published in the reports. This had to be pointed out to NCDIR which was later corrected. This is a missed opportunity as much decision making would have been possible been done on this data.

It is pertinent to note that asbestos exposure, which may be through occupational exposure in a mine, or an asbestos product factory, or through working in a ship dismantling unit, brake lining unit or installation unit or simply through non-occupational exposures such as working in a building having asbestos materials or even as simple as using talcum powder contaminated with asbestos ^3,4,25,26^. The state of mesothelioma in India, especially when asbestos is banned in many countries around the world, needs special focus as large quantities of asbestos are still being imported and used to make asbestos roofing (among other products), is also exported to other developing countries ^27^. It is also possible that workers in marble mines who may otherwise may just be believed to be having silicosis, may develop mesothelioma due to asbestos found in uneconomical quantities in marble quarries and mines. Workers may be unaware as to exposure to asbestos as well ^28^. Another limitation of our study is that though asbestos exposure has been the major cause linked to mesothelioma, there may be instances of mesothelioma from sugarcane farming due to fibres, though a positive correlation has not been found in some studies.^29,30^ Carbon nanotubes are another potential cause ^31^. This does not diminish the fact that the leading cause is asbestos exposure worldwide and is most likely to be the major cause of mesothelioma in India.

This study addresses the issue in previous writings which state that mesothelioma is minimal in India. The results do not support this. The numbers are an alarm bell for this so called ‘rare malignancy’. This study is a positive indicator of the need for good record keeping. Another limitation of this study would be that duplication of recorded cases has not been accounted for, as is suggested for cancer recordkeeping ^32^. This means that a patient who has seen multiple hospitals could be a double entry in this study. This is something that NCDIR database takes into consideration but was not possible in this study as only counts could be provided by hospitals under the RTI Act. But at the same time, the NCDIR may not provide complete information as not all hospitals may be making patient entry based on a common national number equivalent to social security number for easy tracking of these patients.

## Conclusion

This study set out to find out the number of cases of mesothelioma in India. The study found 2213 cases between 2012 till 2022 (and 2023 till date in some hospitals). The study also brought to light the large number of hospitals that remain uncovered under the Indian national cancer registry programme. But the major conclusion from this paper is the large mesothelioma cases go unreported which may mean a large chance of asbestos exposure being recognized in patients having mesothelioma is likely large and alarming. The need for regulating asbestos use or banning it totally, as this preventable disease creates a burden on the healthcare system in India, and may cost much more that the benefit of still using asbestos which is banned in multiple countries around the world. For its use, it must be noted that safer substitutes are available.

It is also recommended that cancer record keeping must be improved in India by firstly, making cancer notifiable in the whole of the country and having population-based registries in every state and every corner of the country. Secondly, the data collection must include national common identity mechanisms so that repeat patients’ data may be easily recognised. Third, the cancer registry programme must become a mandate instead of its being a voluntary process. Fourth, work by states like Tamil Nadu, which have created state-based cancer registries must be lauded and be replicated across the country in all the states and union territories.

## Data Availability

All data produced in the present study are available upon reasonable request to the authors

## Acknowledgements

The authors want to thank the Public Information officers of all the hospitals for facilitating the provision of information. The author wants to thank the Public information officers at NCDIR, NCD-II at Health Ministry, GCRI as well as the Tamil Nadu Cancer Registry project. Many thanks to staff and faculty at Drexel University. Special thanks to Shreya G., Praveen K, Sandeep K for their support.

## Declarations

The first author declares no conflict of interest. The second author regularly participates in medical legal activities regarding asbestos, primarily for plaintiffs.

No external funding has been received for this study.

This study involves no human participants and takes data from the public domain and is exempt from ethics approval.

All data produced in the present study are available upon reasonable request to the authors

